# Association of comorbidities with COVID-19 infection rate and severity: nationwide cohort study with propensity score matching

**DOI:** 10.1101/2021.09.22.21263946

**Authors:** Jiyong Kim, Seong Hun Park, Jong Moon Kim

**Author notes:** Corresponding author: Jong Moon Kim, Address: Department of Rehabilitation Medicine, CHA Bundang Medical Center, CHA University, 59 Yatap-ro, Bundang-gu, Seongnam, Gyeonggi-do, Republic of Korea, 13496.

## Abstract

**Objective:** To describe the association of comorbidities with coronavirus infection-19 (COVID-19) infection rates and severity of infection through Korean nationwide medical system.

**Design:** Nationwide population-based retrospective cohort study.

**Setting:** Korean national health insurance claims database between January 1, 2020, and May 30, 2020.

**Participants:** Patients with positive COVID-19 test and 12 folded controls matched by age, sex and region.

**Main Outcomes Measures:** Outcomes were confirmation of the comorbidities affecting the infection rate and the severity of COVID-19. Patients and outcomes were propensity score matching of factors which may affect COVID-19 infection rate and severity was performed. COVID-19 infections were confirmed through laboratory testing. Severe infection was defined as those who underwent tracheostomy, continuous renal replacement therapy, intensive care unit admission, ventilator use, cardiopulmonary resuscitation, or died.

**Results:** A total of 8070 individuals with positive covid-19 test and 12015 controls were identified. In people aged 60 or older, in those insured with Medicaid, and in the disabled, the proportion corresponding to the severe group of patients showed a tendency to increase. The infection rate of COVID-19 was highest in pulmonary disease (adjusted odds ratio 1.88, 95% confidence interval 1.70 to 2.03), and hyperlipidemia (0.73, 0.67 to 0.80) had a lower infection rate. Disease severity was highest in kidney disease (5.59, 2.48 to 12.63), and lower in hyperlipidemia (0.78, 0.60 to 1.00).

**Conclusions:** There is less bias as the government pays for all tests and treatments related to COVID-19 included in the data used in this study. Using propensity matching to reduce statistical bias, we found that most comorbidities increased the infection rate and severity of COVID-19, whereas hyperlipidemia reduced the rate and severity of infection. These results can be utilized to effectively manage COVID-19 infections.

## INTRODUCTION

Vaccines against coronavirus infection-19 (COVID-19) have been developed, but the number of infections and deaths of COVID-19 is still increasing worldwide.^1^ COVID-19 infection can cause asymptomatic or flu-like symptoms, but some patients are admitted to hospital for conservative treatment, some require ventilator treatment, and in severe cases some patients may die.^2 3^ As the number of people infected with COVID-19 increases, it is important to identify those who are vulnerable to severe COVID-19 infection in order to effectively manage health care facilities.^4 5^

Since the COVID-19 outbreak, many studies have been conducted on demographic factors and comorbidity of infected patients. Most studies have reported similar overall results, but there are differences in detailed results, and some contradictory results have been reported.^6-9^ This difference in results may be due to the diversity of subjects and medical systems, as studies have been conducted in various countries around the world, but it may also be because factors other than COVID-19 infection have not been sufficiently considered.

In this study, the effect of patient comorbidities on the infection rate and severity of COVID-19 was investigated with reduced bias through propensity matching for various variables. We also analyzed demographic factors for patients infected with COVID-19.

## METHODS

### Study design and participants

We conducted a large-scale cohort study using a South Korean national health insurance claims database.^10^ In South Korea, all citizens are registered under the Korean National Health Insurance Service (KNHIS) and this database was used for this study. The KNHIS utilizes a nationwide large scale database system, including information of the diagnostic codes of the International Classification of Diseases (ICD-10), the names of procedures performed, prescription drugs, hospital information, direct medical costs of inpatient and outpatient treatments, as well as information on medical insurance premiums. Since all Koreans are given unique identification numbers at birth and these numbers are used in KNHIS, the health records of patients are not duplicated or omitted.^11 12^ This study used the database ‘‘NHIS-2020-1-328’’ provided by the KNHIS. From January 1 to May 30, 2020, subjects with disease codes B342, B972, U071, U072, MT043, 3/02 were used to identify confirmed COVID-19 patients. A control group equivalent to 15 times the number of confirmed COVID-19 cases with adjusted sex, age, and region was also provided.

### Study population

Laboratory confirmation of COVID-19 infection was defined as a positive result according to World Health Organization guidelines.^13^ We combined the claims-based data from the KNHIS between 1 January 2015 and 30 May 2020 and extracted information on the age, sex and region of residence from the insurance eligibility data. Certain underlying diseases with a high risk of serious illness due to the virus that causes COVID-19, published by the Centers for Disease Control and Prevention and previous studies,^6-9, 14^ were selected; among them, only those with more than 500 people infected with COVID-19 were selected. The history of underlying diseases (Pulmonary disease, Cardiovascular disease, Kidney disease, Hepatobiliary disease, Hyperlipidemia, Gastrointestinal disease, Diabetes mellitus, Hypertension, Psychotic disorder, Dementia, Stroke, Neurogenic disorder, Autoimmune disease, Cancer) was confirmed by the assignment of at least two claims within 1 year using the appropriate ICD-10 code (Supplement 1). The Charlson Comorbidity Index (CCI) score was calculated from the ICD-10 codes by using previously reported methods.^15^

### Outcomes

To determine the severity of the disease according to the demographic factors of COVID-19 infected patients, the severity scale was divided into four grades: mild, moderate, severe, and death. In South Korea, patients with asymptomatic or mild symptoms are discharged when a negative COVID-19 test is confirmed two weeks after hospitalization, which corresponds to the period of self-isolation. When we checked the hospitalization period of COVID-19 infected patients, the hospitalization period peaked on the 16th day and decreased thereafter. Based on this result, the hospitalization period of 16 days or less was defined as a mild grade corresponding to asymptomatic or mild symptoms (Supplement 2-1, 2). Severe grade was defined as tracheostomy, continuous renal replacement therapy, intensive care unit (ICU) admission, ventilator use, and cardiopulmonary resuscitation. Moderate grade was defined as a case where the hospitalization period was longer than 16 days and the treatment corresponding to the severe grade was not performed.

The primary outcome was a comparison of COVID-19 infected and control groups and severity grade according to demographic factors, comorbidity, and complications. The secondary result was performed through propensity score (PS) matching for the comparison target. We identified the degree of infection rate and severity (severe + death / mild + moderate) of COVID-19 according to the comorbidity condition.

### Statistical analysis

We performed PS matching to balance the baseline characteristics of each comorbidity (existence-nonexistence) and to adjust for potential confounders. The PS is estimated using a logistic regression model, calculating the predicted probability of covariates 1) age, 2) CCI (0, 1, or ≥2) was matched for continuous variables, and 3) sex, 4) medication and 5) other comorbidities were matched with binary variables. We assessed each PS matching of the comorbidity existence in a 1:1 ratio using the ‘greedy nearest neighbor’ algorithm and a scale with a caliper of 0.25. Data after PS matching were analyzed by calculating adjusted odds ratios with 95% confidence intervals (CIs) for infection rate and severity (severe + death / mild + moderate) of COVID-19.

## RESULTS

### Clinical characteristics of the study population

A total of 8070 people had positive results on the COVID-19 test. We identified 121,050 uninfected people as control subjects (Figure). The demographic characteristics of the entire cohort are displayed in Table 1. The severity grade of COVID-19 infected persons was mild in 2419 (29.98%), moderate in 5160 (63.94%), severe in 254 (3.15%), and death in 237 (2.94%). Among the total infected people, 3236 (40.10%) were male, and the most common ages were people in their 50s (19.42%) and 60s (14.86%). In terms of the medical insurance grade which indicates socioeconomic status, Medicaid had a high rate of severe or death, but the other grades did not have any obvious tendencies. By region, 5264 (65.23%) of infected people occurred in Daegu, because many infections occurred in Daegu in February and March. People with disabilities had more severe infections and had a much higher mortality rate.

**Figure.**
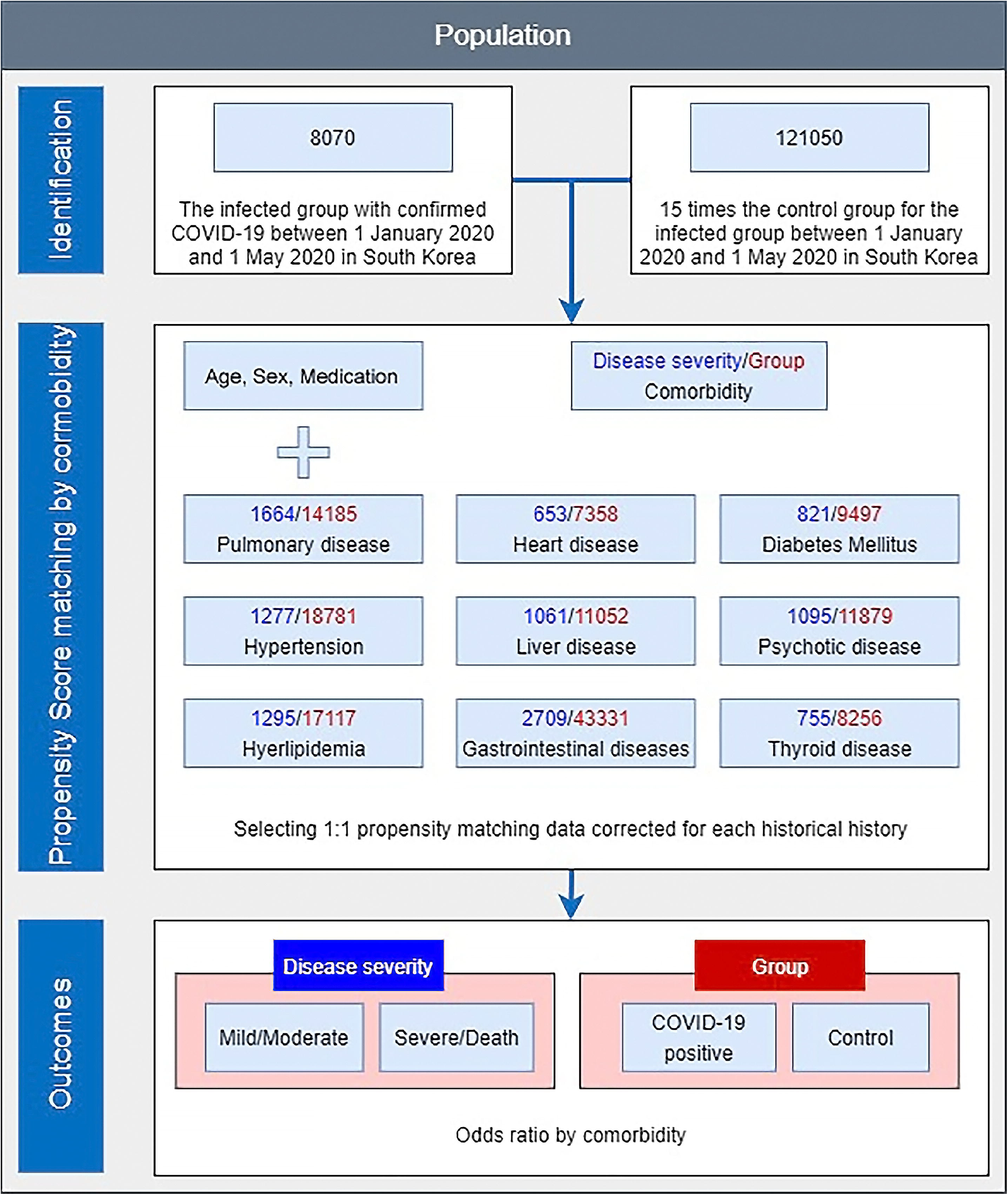
Disposition of patients in the Korean nationwide cohort.

**Table 1.** Baseline characteristics.

| Table 1 \| Baseline characteristics |  |  |  |  |  |  |
| --- | --- | --- | --- | --- | --- | --- |
| Variables | Severity* |  |  |  | COVID-19 | Control§ |
|  | Mild | Moderate | Severe | Death | Total† |  |
| Sex |  |  |  |  |  |  |
| Male | 894 | 2073 | 135 | 134 | 3236 | 48540 |
|  | (27.63) | (64.06) | (4.17) | (4.14) | (40.10) |  |
| Female | 1525 | 3087 | 119 | 103 | 4834 | 72510 |
|  | (31.55) | (63.86) | (2.46) | (2.13) | (59.90) |  |
| Age |  |  |  |  |  |  |
| 0~9 | 32 | 45 | 4 | 0 | 81 | 1215 |
|  | (39.51) | (55.56) | (4.94) | (0.00) | (1.00) |  |
| 10~19 | 77 | 195 | 4 | 0 | 276 | 4140 |
|  | (27.90) | (70.65) | (1.45) | (0.00) | (3.42) |  |
| 20~29 | 697 | 1342 | 18 | 0 | 2057 | 30855 |
|  | (33.88) | (65.24) | (0.88) | (0.00) | (25.49) |  |
| 30~39 | 273 | 541 | 17 | 1 | 832 | 12480 |
|  | (32.81) | (65.02) | (2.04) | (0.12) | (10.31) |  |
| 40~49 | 358 | 655 | 20 | 3 | 1036 | 15540 |
|  | (34.56) | (63.22) | (1.93) | (0.29) | (12.84) |  |
| 50~59 | 504 | 1006 | 43 | 14 | 1567 | 23505 |
|  | (32.16) | (64.20) | (2.74) | (0.89) | (19.42) |  |
| 60~69 | 322 | 776 | 66 | 35 | 1199 | 17985 |
|  | (26.86) | (64.72) | (5.50) | (2.92) | (14.86) |  |
| 70~79 | 122<br>(19.77) | 389<br>(63.05) | 40<br>(6.48) | 66<br>(10.70) | 617<br>(7.65) | 9255 |
| 80~ | 34<br>(8.40) | 211<br>(52.10) | 42<br>(10.37) | 118<br>(29.14) | 405<br>(5.02) | 6075 |
| Medical insurance‡ |  |  |  |  |  |  |
| Medicaid | 186<br>(27.56) | 416<br>(61.63) | 31<br>(4.59) | 42<br>(6.22) | 675<br>(8.36) | 4424<br>(3.65) |
| Grade 1 | 604<br>(32.95) | 1146<br>(62.52) | 44<br>(2.40) | 39<br>(2.13) | 1833<br>(22.71) | 26258<br>(21.69) |
| Grade 2 | 462<br>(30.84) | 971<br>(64.82) | 36<br>(2.40) | 29<br>(1.94) | 1498<br>(18.56) | 24270<br>(20.05) |
| Grade 3 | 484<br>(29.02) | 1081<br>(64.81) | 58<br>(3.48) | 45<br>(2.70) | 1668<br>(20.67) | 27521<br>(22.74) |
| Grade 4 | 637<br>(28.07) | 1475<br>(65.01) | 79<br>(3.48) | 78<br>(3.44) | 2269<br>(28.12) | 37241<br>(30.76) |
| Region |  |  |  |  |  |  |
| Seoul | 102<br>(18.75) | 433<br>(79.60) | 4<br>(0.74) | 5<br>(0.92) | 544<br>(6.74) | 8160 |
| Dae-gu | 1629<br>(30.95) | 3352<br>(63.68) | 133<br>(2.53) | 150<br>(2.85) | 5264<br>(65.23) | 7896 |
| Gyeonggi-do | 156<br>(34.59) | 265<br>(58.76) | 15<br>(3.33) | 15<br>(3.33) | 451<br>(5.59) | 6765 |
| Gyeongsangbuk-do | 266<br>(27.82) | 573<br>(59.94) | 66<br>(6.90) | 51<br>(5.33) | 956<br>(11.85) | 14340 |
| Others | 266<br>(31.11) | 537<br>(62.81) | 36<br>(4.21) | 16<br>(1.87) | 855<br>(10.59) | 12825 |
| Disability grade |  |  |  |  |  |  |
| Mild | 65<br>(20.44) | 192<br>(60.38) | 28<br>(8.81) | 33<br>(10.38) | 318<br>(3.94) | 4367<br>(3.61) |
| Severe | 83<br>(27.57) | 166<br>(55.15) | 18<br>(5.98) | 34<br>(11.30) | 301<br>(3.73) | 2275<br>(1.88) |
| Total | 2419<br>(29.98) | 5160<br>(63.94) | 254<br>(3.15) | 237<br>(2.94) | 8070<br>(100) | 121050<br>(100) |

\*: The number in parentheses means the ratio of each row to the COVID-19 Total
†: The number in parentheses means the ratio of the total number of COVID-19 confirmed cases, 8070 people
§: The number in parentheses means the ratio of 12,1050 total control subjects
‡: Subjects to some specific groups, such as soldiers, are not included.

Those infected with COVID-19 had histories of gastrointestinal disease, pulmonary disease, hyperlipidemia, and hypertension in that order, and the mortality rate was high in cases of kidney disease, dementia, and immunocompromised disease (Table 2-1). After COVID-19 infection was confirmed, gastrointestinal disease, pulmonary disease, and hyperlipidemia were the most common in that order. The mortality rate was high in cases of kidney disease, dementia, and cardiovascular disease (Table 2-2).

**Table 2-1.**
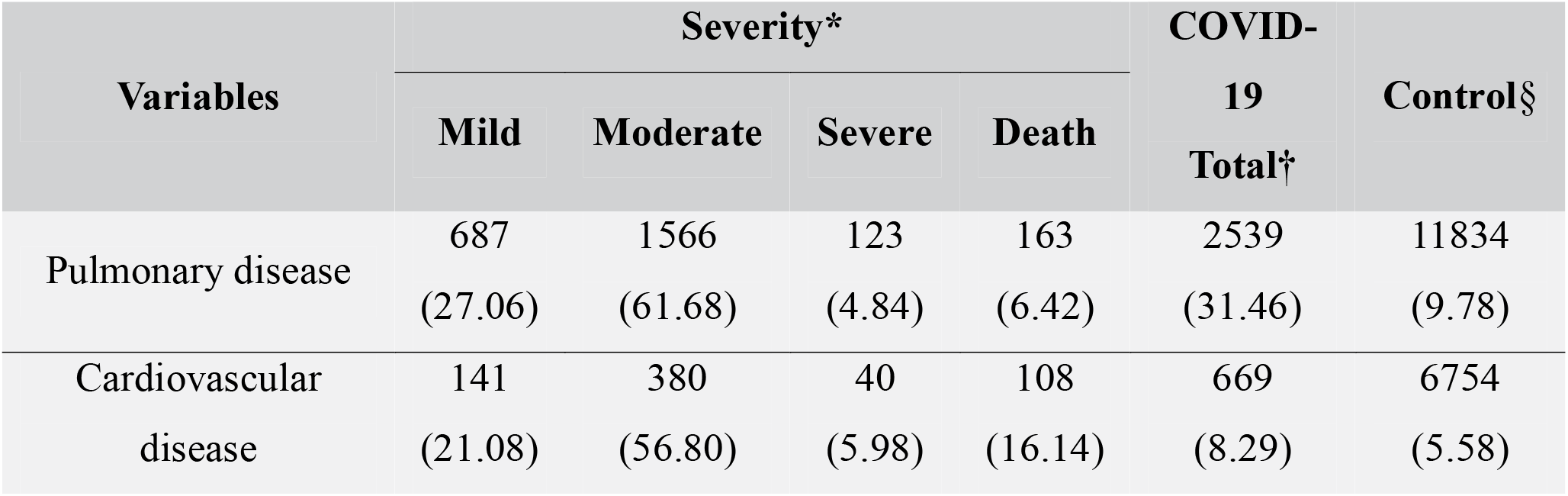
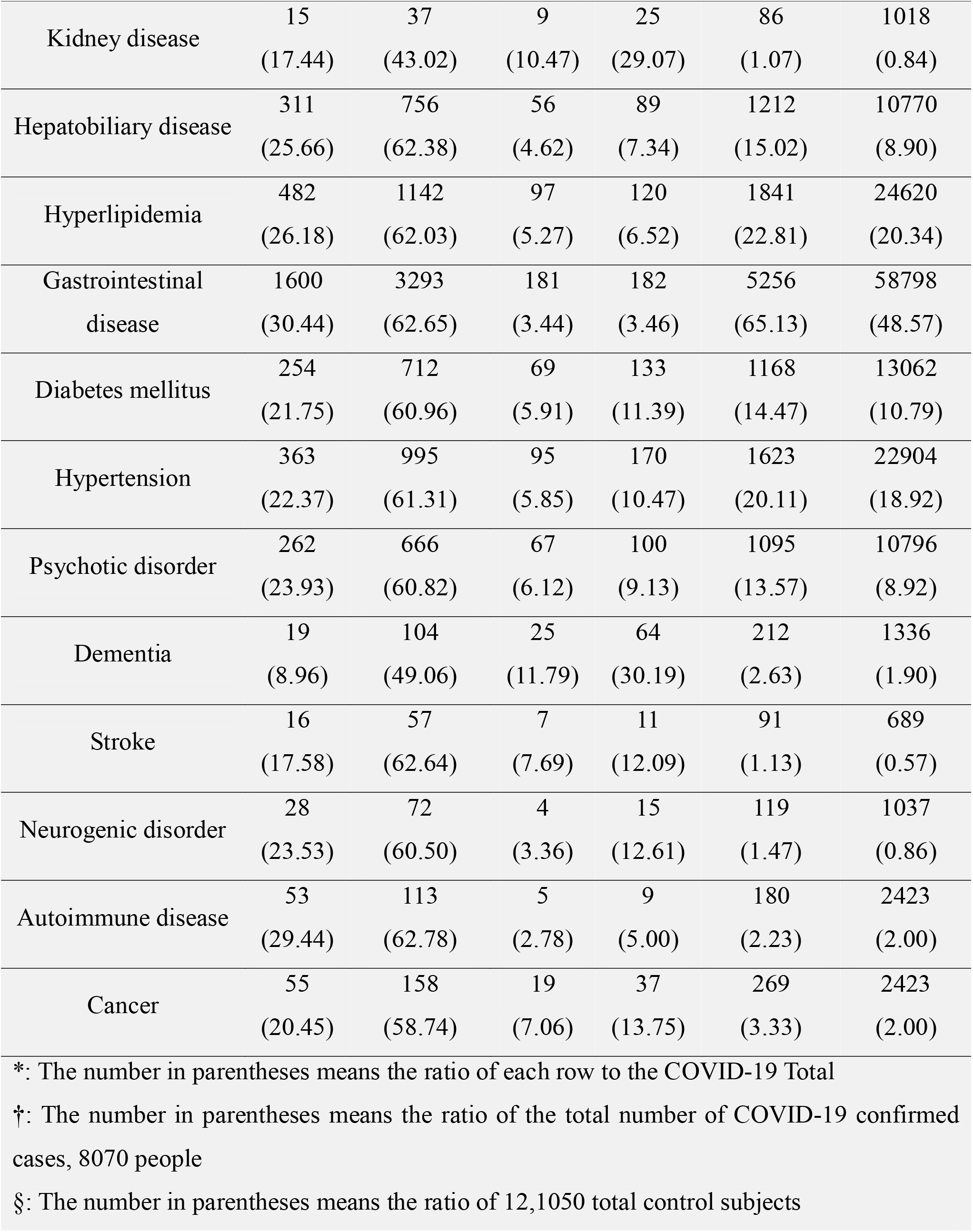
Baseline characteristics of comorbidity.

| Variables | Severity* |  |  |  | COVID-19<br>Total† | Control§ |
| --- | --- | --- | --- | --- | --- | --- |
|  | Mild | Moderate | Severe | Death |  |  |
| Pulmonary disease | 687<br>(27.06) | 1566<br>(61.68) | 123<br>(4.84) | 163<br>(6.42) | 2539<br>(31.46) | 11834<br>(9.78) |
| Cardiovascular<br>disease | 141<br>(21.08) | 380<br>(56.80) | 40<br>(5.98) | 108<br>(16.14) | 669<br>(8.29) | 6754<br>(5.58) |
| Kidney disease | 15<br>(17.44) | 37<br>(43.02) | 9<br>(10.47) | 25<br>(29.07) | 86<br>(1.07) | 1018<br>(0.84) |
| Hepatobiliary disease | 311<br>(25.66) | 756<br>(62.38) | 56<br>(4.62) | 89<br>(7.34) | 1212<br>(15.02) | 10770<br>(8.90) |
| Hyperlipidemia | 482<br>(26.18) | 1142<br>(62.03) | 97<br>(5.27) | 120<br>(6.52) | 1841<br>(22.81) | 24620<br>(20.34) |
| Gastrointestinal<br>disease | 1600<br>(30.44) | 3293<br>(62.65) | 181<br>(3.44) | 182<br>(3.46) | 5256<br>(65.13) | 58798<br>(48.57) |
| Diabetes mellitus | 254<br>(21.75) | 712<br>(60.96) | 69<br>(5.91) | 133<br>(11.39) | 1168<br>(14.47) | 13062<br>(10.79) |
| Hypertension | 363<br>(22.37) | 995<br>(61.31) | 95<br>(5.85) | 170<br>(10.47) | 1623<br>(20.11) | 22904<br>(18.92) |
| Psychotic disorder | 262<br>(23.93) | 666<br>(60.82) | 67<br>(6.12) | 100<br>(9.13) | 1095<br>(13.57) | 10796<br>(8.92) |
| Dementia | 19<br>(8.96) | 104<br>(49.06) | 25<br>(11.79) | 64<br>(30.19) | 212<br>(2.63) | 1336<br>(1.90) |
| Stroke | 16<br>(17.58) | 57<br>(62.64) | 7<br>(7.69) | 11<br>(12.09) | 91<br>(1.13) | 689<br>(0.57) |
| Neurogenic disorder | 28<br>(23.53) | 72<br>(60.50) | 4<br>(3.36) | 15<br>(12.61) | 119<br>(1.47) | 1037<br>(0.86) |
| Autoimmune disease | 53<br>(29.44) | 113<br>(62.78) | 5<br>(2.78) | 9<br>(5.00) | 180<br>(2.23) | 2423<br>(2.00) |
| Cancer | 55<br>(20.45) | 158<br>(58.74) | 19<br>(7.06) | 37<br>(13.75) | 269<br>(3.33) | 2423<br>(2.00) |
| *: The number in parentheses means the ratio of each row to the COVID-19 Total |  |  |  |  |  |  |
| †: The number in parentheses means the ratio of the total number of COVID-19 confirmed cases, 8070 people |  |  |  |  |  |  |
| §: The number in parentheses means the ratio of 12,1050 total control subjects |  |  |  |  |  |  |

**Table 2-2.**
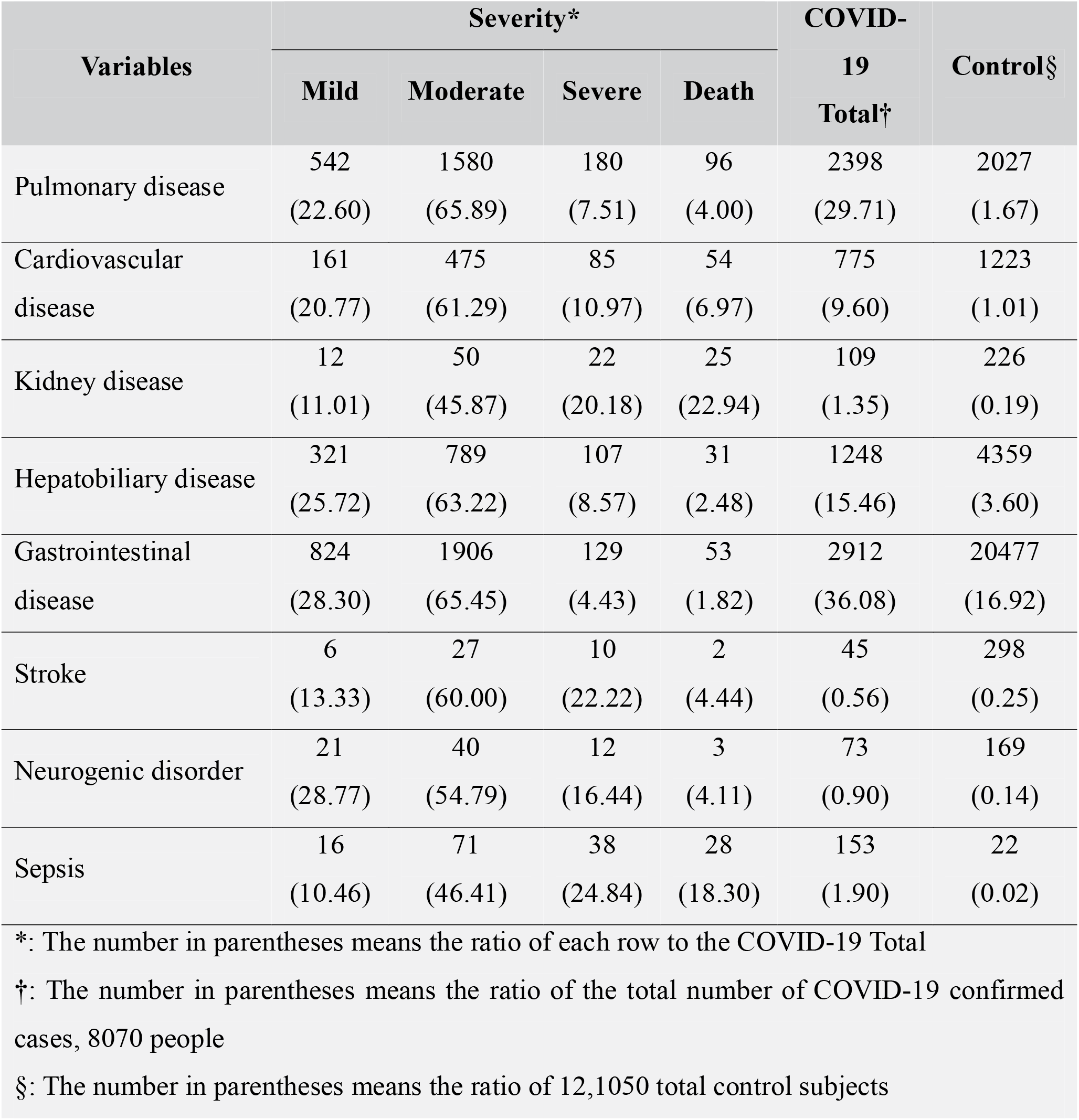
Baseline characteristics of complication.

### COVID-19 test result positivity risks and disease severity according to comorbidity

To identify differences according to comorbidity, predispositions were matched between COVID-19 infected, control group, and COVID-19 infected individuals in each cohort. No significant imbalances in the demographics and clinical characteristics were observed when assessed using standardized mean difference (SMD) within groups of PS-matched cohorts. (The SMD of binary type variables < 0.1) PS-matched odds ratios were checked for age, sex, CCI, comorbidity, and medication. When compared with the control group in the COVID-19 infected group, it was confirmed that COVID-19 infection was more prone for most of the history presented in Table 3, except for the history of hyperlipidemia (adjusted odds ratio, 0.73 [95% CI, 0.67-0.80]), autoimmune disease (0.73, 0.60 to 0.89) and cancer (0.73, 0.62 to 0.86). In COVID-19 infected subjects, the severity grade was high for pulmonary disease (1.72, 1.35 to 2.19), cardiovascular disease (1.54, 1.17 to 2.04), kidney disease (5.59, 2.48 to 12.63), diabetes mellitus (1.43, 1.09 to 1.87), hypertension (1.63, 1.23 to 2.15), psychotic disorder (1.29, 1.01 to 6.52]), dementia (2.92, 1.91 to 4.47), and cancer (1.84, 1.15 to 2.94), and lower with hyperlipidemia (0.70, 0.55 to 0.90) (Table 4).

**Table 3.** Propensity score-matched baseline characteristics and COVID-19 infection test positivity according to comorbidity.

| Variables | Group |  | Adjusted odds ratio<br>(95% CI) |
| --- | --- | --- | --- |
|  | COVID-19 | Control |  |
| Pulmonary disease* | 2880 | 22900 | 1.88 (1.70, 2.03) |
| Cardiovascular disease* | 1245 | 13733 | 1.20 (1.07, 1.35) |
| Kidney disease | 171 | 2037 | 1.01 (0.74, 1.39) |
| Hepatobiliary disease* | 1959 | 20505 | 1.31 (1.19, 1.44) |
| Hyperlipidemia† | 2375 | 25663 | 0.73 (0.67, 0.80) |
| Gastrointestinal disease* | 3164 | 43370 | 1.74 (1.62, 1.88) |
| Diabetes mellitus* | 1544 | 16468 | 1.28 (1.16, 1.43) |
| Hypertension | 1483 | 16473 | 1.04 (0.93, 1.15) |
| Psychotic disorder | 2122 | 21458 | 1.06 (0.97, 1.16) |
| Dementia* | 365 | 2753 | 1.75 (1.40, 2.20) |
| Stroke* | 194 | 1662 | 1.67 (1.23, 2.27) |
| Neurologic disease | 223 | 2089 | 1.16 (0.88, 1.53) |
| Autoimmune disease† | 421 | 4785 | 0.73 (0.60, 0.89) |
| Cancer† | 629 | 6459 | 0.73 (0.62, 0.86) |

\*: Comorbidity to be more susceptible to COVID-19 †: Comorbidity to be less infected with COVID-19

**Table 4.**
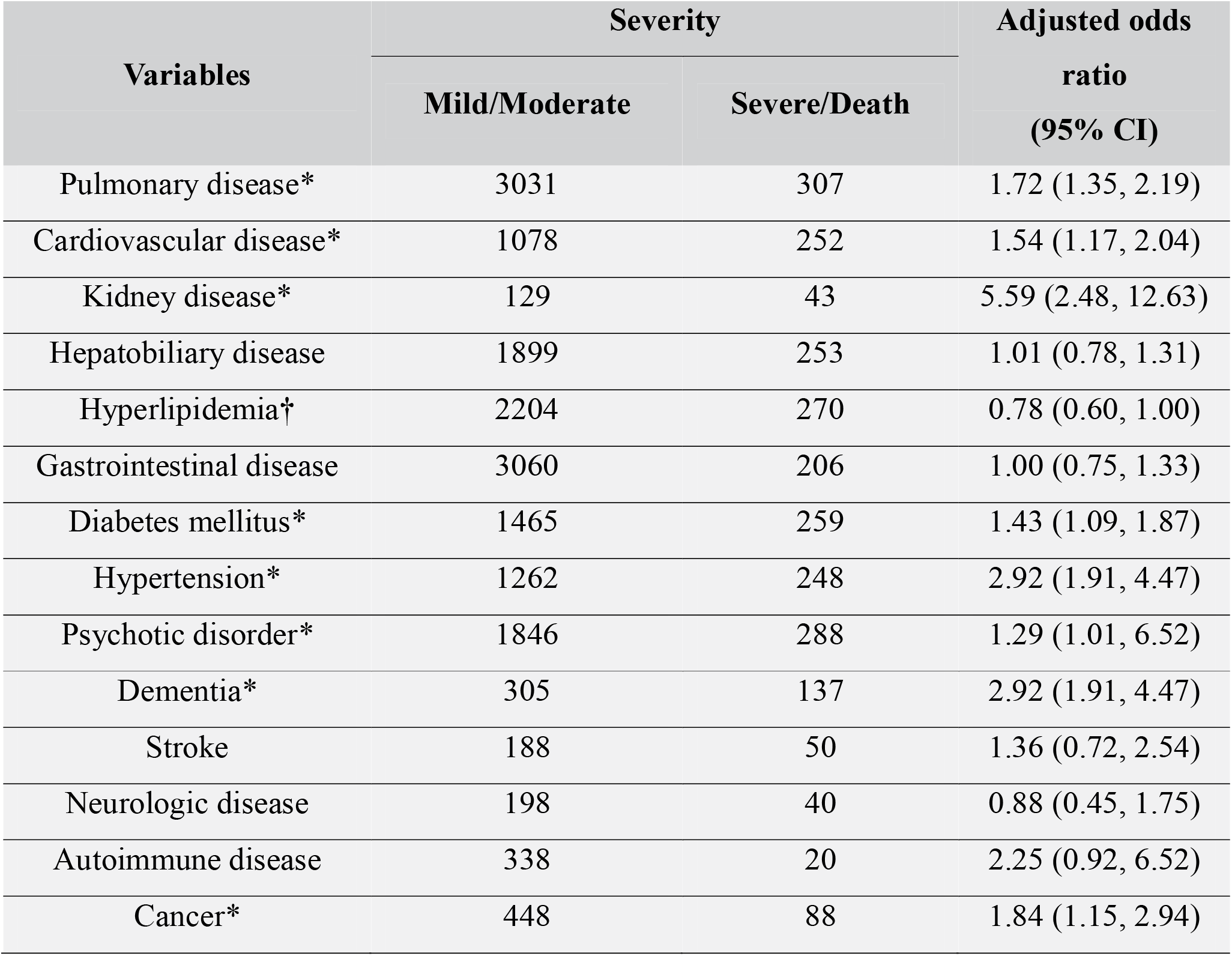
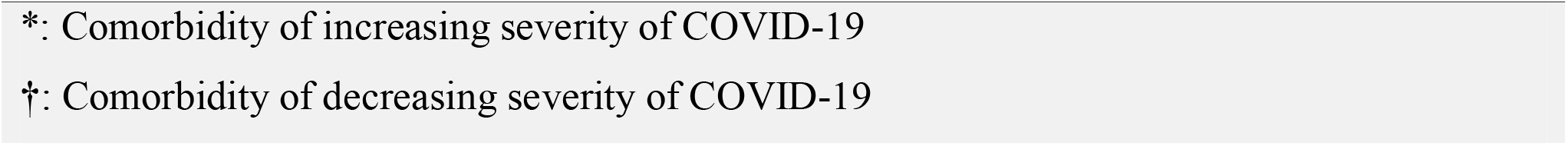
Propensity score-matched baseline characteristics and clinical outcomes of COVID-19 among mild/moderate group versus severe/death groups according to comorbidity in patients with confirmed laboratory COVID-19 infection.

| Variables | Severity |  | Adjusted odds ratio (95% CI) |
| --- | --- | --- | --- |
|  | Mild/Moderate | Severe/Death |  |
| Pulmonary disease* | 3031 | 307 | 1.72 (1.35, 2.19) |
| Cardiovascular disease* | 1078 | 252 | 1.54 (1.17, 2.04) |
| Kidney disease* | 129 | 43 | 5.59 (2.48, 12.63) |
| Hepatobiliary disease | 1899 | 253 | 1.01 (0.78, 1.31) |
| Hyperlipidemia† | 2204 | 270 | 0.78 (0.60, 1.00) |
| Gastrointestinal disease | 3060 | 206 | 1.00 (0.75, 1.33) |
| Diabetes mellitus* | 1465 | 259 | 1.43 (1.09, 1.87) |
| Hypertension* | 1262 | 248 | 2.92 (1.91, 4.47) |
| Psychotic disorder* | 1846 | 288 | 1.29 (1.01, 6.52) |
| Dementia* | 305 | 137 | 2.92 (1.91, 4.47) |
| Stroke | 188 | 50 | 1.36 (0.72, 2.54) |
| Neurologic disease | 198 | 40 | 0.88 (0.45, 1.75) |
| Autoimmune disease | 338 | 20 | 2.25 (0.92, 6.52) |
| Cancer* | 448 | 88 | 1.84 (1.15, 2.94) |

\*: Comorbidity of increasing severity of COVID-19
†: Comorbidity of decreasing severity of COVID-19

## DISCUSSION

This study is a retrospective cohort study conducted in South Korea from January to May 2020 with most of the confirmed COVID-19 infected patients enrolled in medical insurance. Previous studies on the demographic factors of people infected with COVID-19 showed that men, older men, and lower income levels are factors that are more likely to be associated with people infected with COVID-19 and with a higher severity grade.^16 17^ In this study, more women were infected with COVID-19, but the severity grade was higher in men and was directly proportional to age, especially in men over 70 years. In terms of socioeconomic status, the severity grade was higher in the case of Medicaid with the lowest income level, but there was no trend in the rest of the groups. This differs from studies conducted in other countries, possibly because South Korea pays the entire medical cost for COVID-19 infections. In people with disabilities, the incidence was slightly higher than in the control group, but the severity grade was much higher than other people when infected.

According to studies of the COVID-19 virus, the COVID-19 virus binds to the angiotensin-converting enzyme 2 (ACE2) receptor through the viral structural spike protein at the beginning of infection.^18^ ACE2 is expressed to varying degrees in almost all human organs. ACE2 is highly expressed in cardiomyocytes, proximal tubule cells of the kidney, and bladder urinary tract cells, and is abundantly expressed in intestinal cells of the small intestine, especially in the ileum.^18-21^ Therefore, most critically ill patients with COVID-19 infection suffer multiple organ injuries including acute lung injury, acute kidney injury, cardiac injury, hepatobiliary disease, and pneumothorax.^22^

Therefore, to analyze the effect of each comorbidity on the severity grade when infected with COVID-19, it is necessary to consider other comorbidities. In this study, the influence on each comorbidity was confirmed by selecting as many comorbidities as possible for statistically meaningful matching of COVID-19 confirmed patients. Most results were similar to those of previously published studies; however, some results were conflicting.

In this study, people with hyperlipidemia had a low infection rate of COVID-19 and a low severity grade. Previous studies reported that hyperlipidemia should be managed to prevent COVID-19 infection because high cholesterol induces inflammation and increases ACE2 availability.^23-25^ On the other hand, there have been studies showing that patients with hypolipidemia are susceptible to infection, and in some studies, when the lipid level is low, cases of COVID-19 infection are higher than that of the control group, and there are studies showing that the severity level is higher.^26-33^ This may be similar to the ‘obesity paradox’ that describes the fact that it is more advantageous if there is mild obesity in the process of improvement after stroke.^34, 35^ Mild obesity can withstand the systemic catabolic imbalance with impaired metabolic efficiency and degradation of body tissues that occurs after stroke. Hyperlipidemia may also play a role in minimizing the severity of COVID-19 infection. Our study has several limitations. We defined diseases based on ICD codes in insurance claims data. There may be additional unmeasured confounders influencing our results, including genetic polymorphisms, smoking, body mass index and the exposure of the virus. In this study as well, the infection rate of COVID-19 may have been influenced by the degree of exposure to the COVID-19 virus as an important factor in addition to the comorbidity factors.

However, in this study, the influence of COVID-19 itself could be confirmed because the bias was less than that of previous studies. First, South Korea has a population of more than 95% of one race, hence there is minimal racial bias compared to previous studies. Second, because the government funds treatment for COVID-19 infection in South Korea, and the medical facilities for COVID-19 treatment are ubiquitous, there is minimal economic bias. Third, because the PS matching was performed on sex, age, CCI, and comorbidity, selection bias was also minimized. Therefore, more accurate information on the infection rate and disease severity of COVID-19 according to comorbidities could be provided in this study.

In conclusion, certain comorbidities, known as risk factors in previous studies, increase the infection rate and severity of COVID-19; hyperlipidemia decreases the infection rate and severity. These results can be utilized to effectively manage COVID-19 infections.

## Data Availability

The dataset for the national health insurance sharing data is available on request (https://nhiss.nhis.or.kr/bd/ab/bdabd003cv.do).

## Author Contributions

JMK and JK both had full access to all the data in the study and take responsibility for the integrity of the data and the accuracy of the data analysis.

## Concept and design

JMK and JK

## Acquisition, analysis, or interpretation of data

PSH and JMK

## Drafting of the manuscript

JMK and JK

## Statistical analysis

PSH and JK

## Obtained funding

JMK

## Ethical approval

This research protocol was approved by the institutional review board (2020-06-052)

## Data sharing

The dataset for the national health insurance sharing data is available on request (https://nhiss.nhis.or.kr/bd/ab/bdabd003cv.do). Informed consent was not obtained, but the presented data are anonymised and the risk of identification is low.

## Transparency

The lead author (JMK) affirms that the manuscript is an honest, accurate, and transparent account of the study being reported; that no important aspects of the study have been omitted; and that any discrepancies with the study as planned (and, if relevant, registered) have been explained.

## Key points

### WHAT IS ALREADY KNOWN ON THIS TOPIC

In previous studies, studies on comorbidity and the infection rate and severity of COVID-19 were conducted, but conflicting results were also obtained.

### WHAT THIS STUDY ADDS

The relationship between comorbidity and COVID-19 was confirmed by minimizing the bias by using propensity matching and the characteristic that the Korean government pays all costs for COVID-19. While most comorbidities increased the infection rate and severity of COVID-19, hyperlipidemia reduced the infection rate and severity.

