## supplement table for "Association of comorbidities with COVID-19 infection rate and severity: nationwide cohort study with propensity score matching"

| **Supplement 1 \| ICD-10 code of disease used for analysis** | | | | |
| --- | --- | --- | --- | --- |
| **No.** | **Category** | **No.** | **Disease** | **ICD Code** |
| 1 | Pulmonary disease | 1 | Bronchitis | J40.0, J41.0, J41.1, J41.8, J42.0 |
|  |  | 2 | Emphysema | J98.2, J98.3, J43, J43.1, J43.2, J43.8, J43.9, T81.81 |
|  |  | 3 | Chronic obstructive pulmonary disease | J44.0, J44.1, J44.8, J44.9 |
|  |  | 4 | Asthma | J45.0 J45.1, J45.8, J45.9 |
|  |  | 5 | Bronchiectasis | J47.0 |
|  |  | 6 | Pulmonary eosinophilia | J82.0, J82.8 |
|  |  | 7 | Interstitial pulmonary disease | J84.0, J84.1, J84.8, J84.9 |
|  |  | 8 | Pulmonary Tb | A15.0, A15.1, A15.2, A15.3, A15.4, A15.5, A15.6, A15.7, A15.8, A15.9, A16.0, A16.1, A16.2, A16.3, A16.4, A16.5, A16.7, A16.8, A16.9, U84.3 |
| 2 | Cardiovascular disease | 1 | Angina Pectoris | I20.0, I2.1, I20.8, I20.9 |
|  |  | 2 | Myocardial infarction | I21.0, I21.1, I21.2, I21.3, I21.4, I21.9, I22.0, I22.1, I22.8, I22.9 |
|  |  | 3 | Ischemic heart disease | I24.0, I24.1, I24.8, I24.9, I25.0, I25.1, I25.2, I25.3, I25.4, I25.5, I25.6, I25.8, I25.9 |
|  |  | 4 | Cardiomyopathy | I25.5, I42.0, I42.1, I42.2, I42.3, I42.4, I42.5, I42.6, I42.7, I42.8, I42.9, I43.0, I43.1, I43.2, I43.8 |
|  |  | 5 | Arrhythmia | I44.0, I44.1, I44.2, I44.3, I44.4, I44.5, I44.6, I44.7, I45.0, I45.1, I45.2, I45.3, I45.4, I45.5, I45.6, I45.8, I45.9, I46.0, I46.1, I46.9, I47.0, I47.1, I47.2, I47.9, I49.0, I49.1, I49.2, I49.3, I49.4, I49.5, I49.8, I49.9 |
|  |  | 6 | Atrial fibrillation | I48.0, I48.1, I48.2, I48.3, I48.4, I48.9 |
|  |  | 7 | Heart failure | I50.0, I50.1, I50.9 |
|  |  | 8 | Atherosclerosis | I70.0, I70.1, I70.2, I70.8, I70.9 |
|  |  | 9 | thromboembolism | I74.0, I74.1, I74.2, I74.3, I74.4, I74.5, I74.8, I74.9, I80.0, I80.1, I80.2, I80.3, I80.8, I80.9, I81, I82.0, I82.1, I82.2, I82.3, I82.8, I82.9, I26.0, I26.9 |
| 3 | Kidney disease | 1 | Chronic kidney disease | N18.1, N18.2, N18.3, N18.4, N18.5, N18.9 |
|  |  | 2 | Unspecified kidney failure | N19 |
| 4 | Hepatobiliary disease | 1 | Hepatic failure | K72.0, K72.1, K72.9 |
|  |  | 2 | Chronic hepatitis | K73.0, K73.1, K73.2, K73.8, K73.9 |
|  |  | 3 | Fibrosis and cirrhosis of liver | K74.0, K74.1, K74.2, K74.3, K74.4, K74.5, K74.6 |
|  |  | 4 | Chronic viral hepatitis | B18.0, B18.1, B18.2, B18.8, B18.9 |
| 5 | Hyperlipidemia | 1 | Disorders of lipoprotein metabolism and other lipidaemia | E78.0, E78.1, E78.2, E78.3, E78.4, E78.5, E78.6, E78.8, E78.9 |
| 6 | Gastrointestinal disease | 1 | Gastrointestinal disease | K21.0, K21.9, K25.0, K25.1, K25.2, K25. 3, K25.4, K25.5, K25.6, K25.7, K25.9, K26.0, K26.1, K26.2, K26.3, K26.4, K26.5, K26.6, K26.7, K26.9, K27.0, K27.1, K27.2, K27.3, K27.4, K27.5, K27.6, K27.7, K27.9, K28.0, K28.1, K28.2, K28.3, K28.4, K28.5, K28.6, K28.7, K28.9, K29.0, K29.1, K29.2, K29.3, K29.4, K29.5, K29.6, K29.7, K29.8, K29.9, K50.0, K50.1, K50.8, K50.9, K51.0, K51.2, K51.3, K51.4, K51.5, K51.8, K51.9, K90.0, K90.1, K90.2, K90.3, K90.4, K90.8, K90.9 |
| 7 | Diabetes Mellitus | 1 | Type 1 diabetes mellitus | E10.0, E10.1, E10.2, E10.3, E10.4, E10.5, E10.6, E10.7, E10.8, E10.9 |
|  |  | 2 | Type 2 diabetes mellitus | E11.0, E11.1, E11.2, E11.3, E11.4, E11.5, E11.6, E11.7, E11.8, E11.9 |
|  |  | 3 | Other specified diabetes mellitus | E13.0, E13.1, E13.2, E13.3, E13.4, E13.5, E13.6, E13.7, E13.8, E13.9 |
|  |  | 4 | Unspecified diabetes mellitus | E14.0, E14.1, E14.2, E14.3, E14.4, E14.5, E14.6, E14.7, E14.8, E14.9 |
| 8 | Hypertension | 1 | Hypertensive disease | I10.1, I10.9, I11.0, I11.9, I12.0, I12.9, I13.0, I13.1, I13.2, I13.9, I15.0, I15.1, I15.2, I15.8, I15.9. |
| 9 | Psychotic disorder | 1 | Organic, including symptomatic, mental disorders except dementia | F04.0, F05.0, F05.1, F05.8, F05.9, F06.0, F06.1, F06.2, F06.3, F06.4, F06.5, F06.6, F06.7, F06.8, F06.9, F07.0, F07.1, F07.2, F07.8, F07.9, F09.0, F20.0, F20.1, F20.2, F20.3, F20.4, F20.5, F20.6, F20.8, F20.9, F21.0, F90.0, F90.1, F90.8, F90.9, F91.0, F91.1, F91.2, F91.3, F91.8, F91.9 |
|  |  | 2 | Depression | F32.0, F32.1, F32.2, F32.3, F32.8, F32.9, F33.0, F33.1, F33.2, F33.3, F33.4, F33.8, F33.9 |
|  |  | 3 | Anxiety, Adjustment disorders | F40.0, F40.1, F40.2, F40.8, F40.9, F41.0, F41.1, F41.2, F41.3, F41.8, F41.9, F43.0, F43.1, F43.2, F43.8, F43.9 |
| 10 | Dementia | 1 | Dementia in Alzheimer disease | F00.0, F00.1, F00.2, F00.9 |
|  |  | 2 | Vascular dementia | F01.0, F01.1, F01.2, F01.3, F01.8, F01.9 |
|  |  | 3 | Dementia in other diseases classified elsewhere | F02.0, F02.1, F02.2, F02.3, F02.4, F02.8 |
|  |  | 4 | Unspecified dementia | F03.0 |
| 11 | Stroke | 1 | Subarachnoid haemorrhage | I60.0, I60.1, I60.2, I60.3, I60.4, I60.5, I60.6, I60.7, I60.8, I60.9 |
|  |  | 2 | Intracerebral haemorrhage | I61.0, I61.1, I61.2, I61.3, I61.4, I61.5, I61.6, I61.8, I61.9 |
|  |  | 3 | Other nontraumatic intracranial haemorrhage | I62.0, I62.1, I62.9 |
|  |  | 4 | Cerebral infarction | I63.0, I63.1, I63.2, I63.3, I63.4, I63.5, I63.6, I63.8, I63.9 |
|  |  | 5 | Stroke, not specified as haemorrhage or infarction | I64.0 |
|  |  | 6 | Occlusion and stenosis of precerebral and cerebral arteries, not resulting in cerebral infarction | I65.0, I65.1, I65.2, I65.3, I65.8, I65.9, I66.0, I66.1, I66.2, I66.3, I66.4, I66.8, I66.9 |
|  |  | 7 | Other cerebrovascular diseases, Cerebrovascular disorders in diseases classified elsewhere | I67.0, I67.1, I67.2, I67.3, I67.4, I67.5, I67.6, I67.7, I67.8, I67.9, I68.0, I68.1, I68.2, I68.8 |
|  |  | 8 | Sequelae of cerebrovascular disease | I69.0, I69.1, I69.2, I69.3, I69.4, I69.8 |
| 12 | Neurologic disease | 1 | Epilepsy, Status epilepticus | G40.0, G40.1, G40.2, G40.3, G40.4, G40.5, G40.6, G40.7, G40.8, G40.9, G41.0, G41.1, G41.2, G41.8, G41.9 |
|  |  | 2 | Encephalitis, myelitis, and encephalomyelitis | G04.0, G04.1, G04.2, G04.8, G04.9, G05.0, G05.1, G05.2, G05.8 |
|  |  | 3 | Demyelinating diseases of the central nervous system | G35.0, G36.0, G36.1, G36.8, G36.9, G37.0, G37.1, G37.2, G37.3, G37.4, G37.5, G37.8, G37.9, G61.0 |
|  |  | 4 | Myasthenia gravis | G70.0 |
| 13 | Autoimmune disease | 1 | Graves’ disease | E05.0 |
|  |  | 2 | Hashimoto thyroiditis | E06.3 |
|  |  | 3 | Rheumatoid arthritis | M05.0, M05.1, M05.2, M05.3, M05.8, M05.9, M06.0, M06.1, M06.2, M06.3, M06.4, M06.8, M06.9 |
|  |  | 4 | Systemic lupus erythematous | M32.0, M32.1, M32.8, M32.9 |
|  |  | 5 | Crohn’s disease | K50.0, K50.1, K50.8, K50.9 |
|  |  | 6 | Mucocutaneous lymph node syndrome [Kawasaki] | M30.3 |
| 14 | Cancer | 1 | Malignant neoplasms | C code (C00-C97) |

| **Supplement 2-1** \| **COVID-19 infection test positivity odds ratios according to different propensity score matching** | | | |
| --- | --- | --- | --- |
| **Variables** | **Adjusted odds ratio (95% CI)** | | |
|  | **PSM 1** | **PSM 2** | **PSM 3** |
| Pulmonary disease | -* | 2.53 (2.33, 2.74) | 1.88 (1.70, 2.03) |
| Cardiovascular disease | 0.44 (0.40, 0.48) | 0.96 (0.86, 1.07) | 1.20 (1.07, 1.35) |
| Kidney disease | -* | 0.94 (0.69, 1.28) | 1.01 (0.74, 1.39) |
| Hepatobiliary disease | -* | 1.49 (1.35, 1.64) | 1.31 (1.19, 1.44) |
| Hyperlipidemia | 0.61 (0.57, 0.65) | 0.95 (0.87, 1.03) | 0.73 (0.67, 0.80) |
| Gastrointestinal disease | 1.71 (1.62, 1.81) | 1.49 (1.41, 1.57) | 1.74 (1.62, 1.88) |
| Diabetes mellitus | 1.12 (1.02, 1.22) | 1.22 (1.12, 1.34) | 1.28 (1.16, 1.43) |
| Hypertension | 1.52 (1.40, 1.64) | 0.92 (0.85, 1.00) | 1.04 (0.93, 1.15) |
| Psychotic disorder | -* | 1.52 (1.38, 1.67) | 1.06 (0.97, 1.16) |
| Dementia | -* | 2.30 (1.84, 2.88) | 1.75 (1.40, 2.20) |
| Stroke | -* | 1.56 (1.16, 2.09) | 1.67 (1.23, 2.27) |
| Neurologic disease | -* | 1.50 (1.12, 2.01) | 1.16 (0.88, 1.53) |
| Autoimmune disease | -* | 1.05 (0.85, 1.30) | 0.73 (0.60, 0.89) |
| Cancer | -* | 0.44 (0.38, 0.51) | 0.73 (0.62, 0.86) |
| *: No matching results due to inadequate covariates  PSM 1: Sex, Age and Charlson Comorbidity Index  PSM 2: PSM 1 + comorbidities associated with COVID-19 infection  PSM 3: PSM 2 + Drugs related to COVID-19 infection | | | |

| **Supplement 2-2** \| **COVID-19 severity odds ratios according to different propensity score matching** | | | |
| --- | --- | --- | --- |
| **Variables** | **Adjusted odds ratio (95% CI)** | | |
|  | **PSM 1** | **PSM 2** | **PSM 3** |
| Pulmonary disease | 1.61 (1.28, 2.03) | 1.48 (1.18, 1.86) | 1.72 (1.35, 2.19) |
| Cardiovascular disease | 2.68 (1.96, 3.67) | 1.57 (1.19, 2.08) | 1.54 (1.17, 2.04) |
| Kidney disease | 4.97 (2.26, 10.93) | 3.09 (1.53, 6.26) | 5.59 (2.48, 12.63) |
| Hepatobiliary disease | 1.50 (1.13, 1.98) | 1.04 (0.80, 1.35) | 1.01(0.78, 1.31) |
| Hyperlipidemia | 1.21 (0.98, 1.49) | 0.92 (0.72, 1.17) | 0.78 (0.60, 1.00) |
| Gastrointestinal disease | 1.02 (0.79, 1.31) | 1.06 (0.82, 1.38) | 1.00 (0.75, 1.33) |
| Diabetes mellitus | 1.86(1.46, 2.37) | 1.44 (1.14, 1.83) | 1.43 (1.09, 1.87) |
| Hypertension | 2.02 (1.62, 2.52) | 1.64 (1.28, 2.10) | 1.63 (1.23, 2.15) |
| Psychotic disorder | 2.09 (1.59, 2.74) | 1.61 (1.25, 2.08) | 1.29 (1.01, 1.66) |
| Dementia | 6.51 (3.97, 10.68) | 3.51 (2.29, 5.36) | 2.92 (1.91, 4.47) |
| Stroke | 2.13 (1.07, 4.24) | 1.55 (0.81, 2.97) | 1.36 (0.72, 2.54) |
| Neurologic disease | 1.87 (0.85, 4.11) | 0.75 (0.39, 1.46) | 0.89 (0.45, 1.75) |
| Autoimmune disease | 1.43 (0.62, 3.32) | 1.08 (0.49, 2.38) | 2.45 (0.92, 6.52) |
| Cancer | 3.27 (1.90, 5.63) | 1.60 (1.02, 2.51) | 1.84 (1.15, 2.94) |
| PSM 1: Sex, Age and Charlson Comorbidity Index  PSM 2: PSM 1 + comorbidities associated with COVID-19 infection  PSM 3: PSM 2 + Drugs related to COVID-19 infection | | | |
